# Uptake and outcomes of earlier viral load testing and longer visit intervals in the early HIV treatment period: retrospective cohort analysis of South Africa’s 2023 ART guidelines revision

**DOI:** 10.64898/2026.11.15.26368143

**Authors:** E Kachingwe, M Benade, A Morgan, N Scott, M Maskew, V Ntjikelane, L Sande, N Mutanda, M Manganye, L Malala, S Rosen

## Abstract

South Africa’s 2023 revised ART Guidelines moved the first scheduled viral load (VL) test from 6 to 3 months after antiretroviral therapy (ART) initiation and accelerated differentiated care eligibility from 7 to 4 months among virally suppressed clients. We assessed uptake of these changes and their association with virologic outcomes.

We analysed routine electronic medical record data from 18 primary healthcare clinics in three South African districts among all adults initiating or re-initiating ART (N=16,338; January 2022-March 2025). Interrupted time series analyses were used to assess uptake of 3-month VL testing and assignment to ≥6-month visit intervals following guideline implementation in April 2023. Modified Poisson regression was used to compare relative risks (RRs) for viral suppression at the first and second routine VL tests among clients tested at 3 months (test window=56-111 days) and 6 months (112-<252 days).

At the first VL assessment, 27% of participants had VL tests at 3 months and 30% at 6 months; the rest had tests outside either window (19%) or no test documented (25%). Three-month VL testing rose immediately after guideline implementation, reaching 72-85% coverage by the end of the study period. Assignment to ≥6-month visit intervals among clients with 3-month viral suppression increased after implementation, reaching 48-71% across districts. A second VL result was documented for 62% of clients tested in the 3- or 6-month window. Viral suppression did not differ meaningfully between clients initially tested at 3 months (72%) and those initially tested at 6 months (72%). Among clients with a second scheduled VL result, 82% were virally suppressed. Assignment to ≥6-month visit intervals was not associated with subsequent viral suppression (aRR 1.02, 95% CI 0.99-1.06). Results should be interpreted with caution due to missing VL data.

South Africa’s 2023 guideline revision was associated with rapid increases in 3-month VL testing and assignment to longer visit intervals. Viral suppression at the first VL test was similar among clients tested at 3 and 6 months, and longer visit intervals were not associated with poorer outcomes. These findings support earlier differentiation of HIV care within routine service delivery. Incomplete uptake of recommended testing and visit intervals offers opportunities to strengthen implementation and warrants continued evaluation of longer-term outcomes.

## INTRODUCTION

Despite the tremendous progress of sub-Saharan Africa’s HIV treatment programs in achieving very high levels of coverage with antiretroviral therapy (ART)(1), disengagement from care remains a major challenge to reducing HIV-related morbidity and mortality, particularly during the first year after treatment initiation or re-initiation(2–4). In South Africa, home to the world’s largest HIV treatment program, a recent analysis of routine medical record data from three provinces found that approximately one in six (16%) clients disengaged from care within six months of initiation, and only 59% remained continuously in care, with no treatment interruptions of greater than 28 days, during the first six months(5). At 12 months, the corresponding figures were 23% and 45%. Other countries, such as Zambia(6) and Malawi(7), face similar levels of disengagement and interruption.

Over the past decade, many countries in the region have implemented policy and guideline changes designed to improve retention in care and reduce barriers to HIV treatment. These include the introduction of differentiated service delivery (DSD) models (called differentiated models of care (DMOCs) in South Africa) and same-day ART initiation (SDI). SDI has effectively eliminated loss to follow up between HIV testing and ART initiation for those who wish to start treatment (8), while DSD models such as community-based medication collection and multi-month dispensing intervals have reduced the burden of treatment on those who are “established” in care(9), typically defined as having at least 6 months’ experience on ART and a suppressed viral load (VL).

While these policies successfully streamlined care for newly initiating and established clients(10,11), only recently have changes been made that benefit those in the period between initiation and DMOC eligibility. The first six months after treatment initiation, known as the “early treatment period,” have traditionally been characterized by frequent clinic visits and heightened risk of disengagement. South Africa began to address this gap in 2023 with its revised Adherence Guidelines for HIV, TB and NCDs(12). The new guidelines moved the first VL test to three months after initiation rather than six, made those with viral suppression at three months eligible for DMOC enrolment at four months rather than seven, and reduced the number of clinic visits required during the first year on treatment. Taken together, these changes comprise a novel model of service for the early treatment period, a longstanding goal for service delivery innovation(13).

Although implementation began in 2023, little is known about the consequences of South Africa’s guideline revisions for treatment outcomes. In particular, it is unclear the extent to which facilities have adopted the three central changes most salient to the early treatment period—early VL testing, earlier enrolment in DMOCs, and fewer clinic visits—and whether these changes are associated with changes in viral suppression. To assess this gap, we evaluated fidelity to and outcomes of these changes in the 2023 South African ART Guidelines.

## METHODS

We analysed routinely collected electronic medical record data to assess uptake of key guideline components and associated clinical outcomes.

### Study sites and population

The study was conducted at 18 public sector primary healthcare clinics (PHCs) in three South African provinces. Six facilities were located in each of King Cetshwayo District (KC) in KwaZulu-Natal, Ehlanzeni District (EH) in Mpumalanga, and West Rand District (WR) in Gauteng. All of these sites are part of the SENTINEL network, originally created in 2020 to monitor DMOC uptake and outcomes (14). Sites were selected to capture both urban and rural settings, the range of differentiated models of care being offered at that time, and assistance from diverse nongovernmental partner organizations supported by the U.S. President’s Emergency Plan for AIDS Relief (PEPFAR). The sites and the clients they serve have been described in several previous publications(14–18).

Adult (≥18 years) clients who initiated or re-initiated ART at one of the study sites from January 1, 2022 onward were eligible for inclusion. We excluded pregnant women, for whom treatment guidelines differ, but imposed no other restrictions at cohort entry. Eligibility for the analytic cohort was subsequently restricted to clients with sufficient follow-up time to reach the final scheduled VL testing window.

### Data collection

Data were extracted from TIER.Net, South Africa’s electronic medical record system for HIV treatment. Variables extracted included demographic characteristics, ART regimens, ART initiation date, clinic visit dates, VL test dates and results, appointment dates, and prescription information. Data were available from January 1, 2022 to April 30, 2026 for King Cetshwayo and Ehlanzeni districts and from January 1, 2022 to November 30, 2025 for West Rand District and were accessed on June 9 2026 for King Cetshwayo, May 9, 2026 for Ehlanzeni, and July 15, 2026 for West Rand. For each patient, available follow-up time was determined individually based on their ART initiation date and the end of the available observation period. The analytic cohort was restricted to patients with sufficient follow-up to reach the final scheduled VL testing window (at least 392 days).

TIER.Net includes a field for recording enrolment in DMOCs, but completion of this field was incomplete in the study dataset. Recorded DMOC enrolment in the TIER.Net study dataset was substantially lower than estimates reported through routine programmatic monitoring and lower than levels observed in previous studies conducted at the same facilities. We therefore used assignment to six-month clinic visits as a proxy measure of differentiated service delivery uptake. Because clients may receive extended visit intervals without formal DMOC enrolment, this proxy measure may overestimate true DMOC enrolment, although it should provide a reasonable reflection of eligibility for differentiated service delivery at the four-month timepoint.

### Intervention description

For this study, we focused on adherence to three core changes introduced in the 2023 guideline revision: 1) shifting of the first scheduled VL test from six months to three months after ART initiation; 2) shifting of eligibility assessment for six-month dispensing (3MMD), six-monthly clinic visits, and enrolment in a DMOC from seven months to four months after initiation (among clients who were virally suppressed); and 3) reduction in the recommended number of clinic visits in the first year from 11 to 6. These changes are illustrated in Figure 1. We note that the second scheduled VL test is recommended at 10 months after initiation for those whose first test is at 3 months and at 12 months for those whose first test is at 6 months(19).

**Figure 1.**
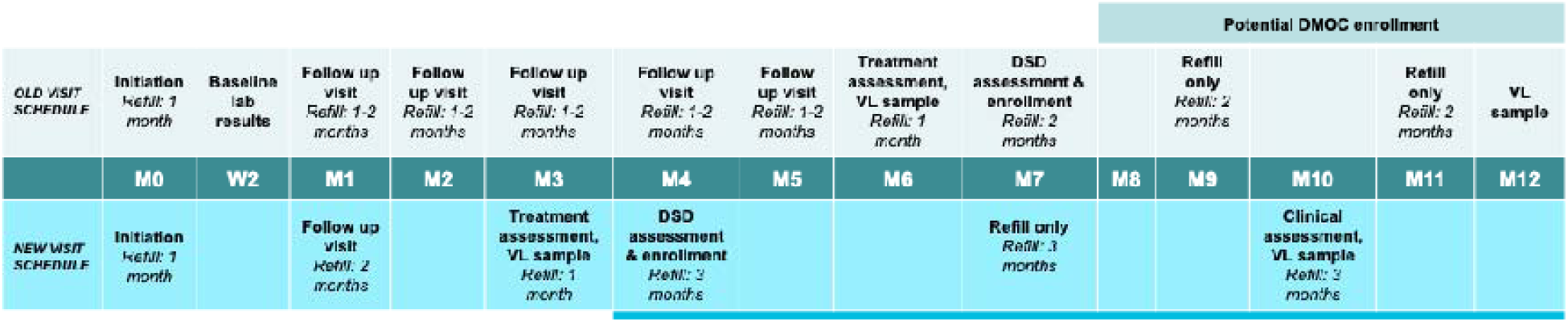
Comparison of old and new treatment schedules for the first year of ART in South Africa.

### Fidelity indicators and clinical outcomes

As measures of guideline fidelity, we assessed adherence to each of the three core components mentioned above: (1) uptake of VL testing approximately three months after ART initiation, defined as VL test observed in the EMR between two and four months after ART initiation; (2) assignment of virally suppressed clients to six-month clinic visit intervals as a proxy for DMOC enrolment; and (3) reduction in the number of clinic visits during the first year on ART, using a count of the number of clinic visits recorded during the first year after ART initiation. Table 1 defines study outcomes, and results are presented in the order of Table 1. These outcomes are also illustrated in Figure 2, showing the steps in the cascade of care proposed by the new guidelines. Note that one dispensing cycle was defined as 28 days, consistent with the South African 2023 ART Clinical Guidelines(20).

**Figure 2.**
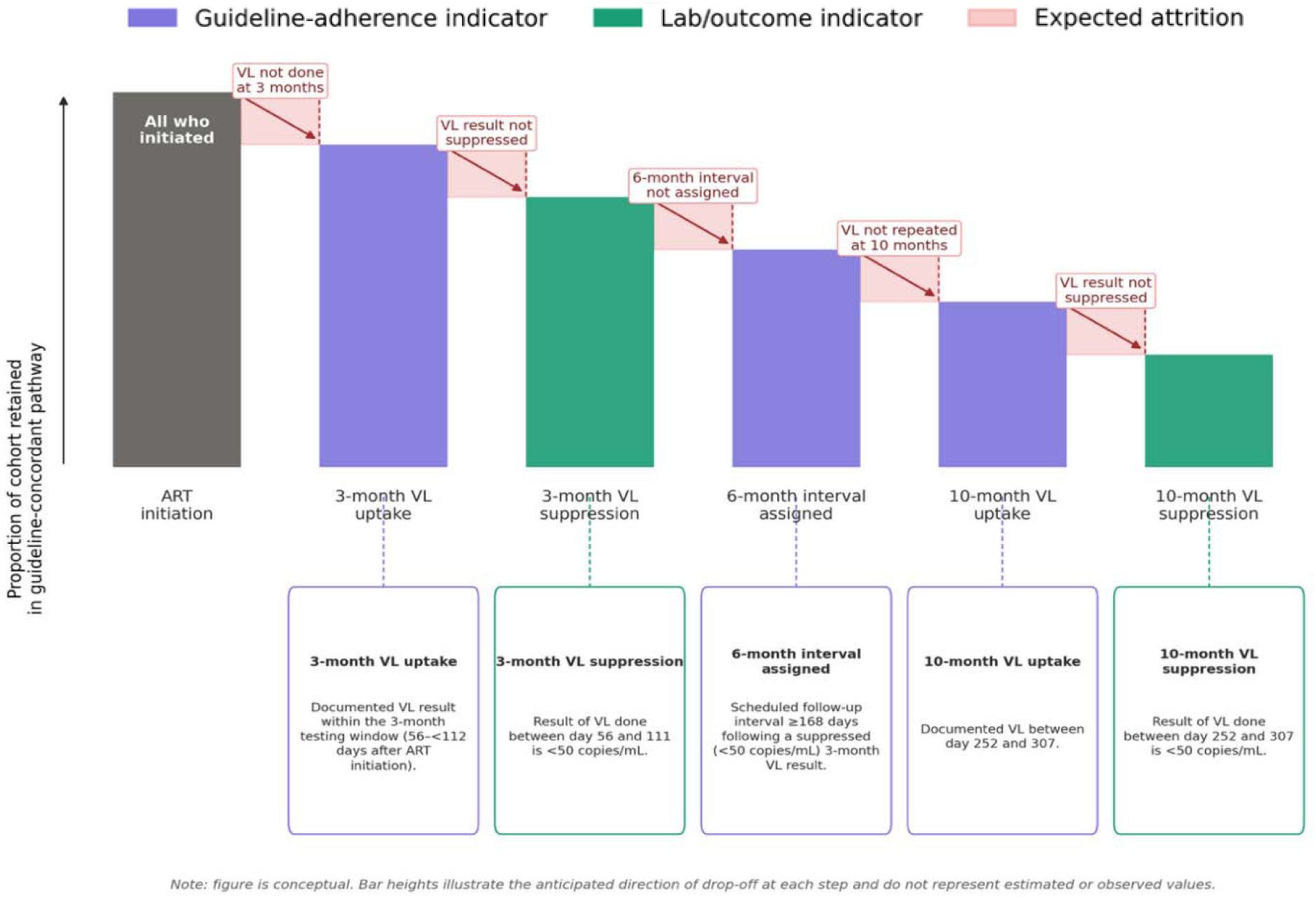
Anticipated care cascade for clients receiving 3-month viral load monitoring under 2023 guidelines.

**Table 1.** Study outcomes and operational definitions.

| Category | Metric or outcome | Definition | Numerator | Denominator |
|---|---|---|---|---|
| *Guideline changes* |  |  |  |  |
| Shifting the first scheduled VL test from 6 months to 3 months after ART initiation | Uptake of 3-month VL testing | Documented VL result within the 3-month testing window (56–111 days after ART initiation) | Clients with a documented VL result within 56–11 days after ART initiation | Study-eligible ART initiators and re-initiators (clients recorded in TIER.Net as initiating ART) |
| Shifting eligibility for differentiated care to month 4 after ART initiation | Uptake of ≥6-month visit intervals following viral suppression | Scheduled follow-up interval ≥168 days following a suppressed (<50 copies/mL) 3-month VL result | Clients assigned a scheduled follow-up interval ≥168 days following a suppressed first VL result | Clients with a suppressed first VL |
| Shifting the second scheduled VL test from 12 months to 10 months after ART initiation | Uptake of second scheduled VL test | Documented VL result within the applicable second scheduled VL testing window: 10 months (window 252-307 days) or 12 months (window 308-39 days) after ART initiation | Clients with a documented VL result within the applicable 10- or 12-month testing window | All clients in the analytic cohort with sufficient follow-up (≥392 days after ART initiation) to reach the second scheduled VL assessment. Uptake among clients with a documented first scheduled VL result was additionally assessed using the subgroup tested within the 3- or 6-month window. |
| Reducing clinic visit frequency during the first year on ART | Clinic visit frequency during the first 12 months of ART | Number of recorded clinic visits within 12 months after ART initiation | N/A (count) | Study-eligible ART initiators retained in care throughout the first 12 months after ART initiation |
| *Treatment outcomes* |  |  |  |  |
| Early viral suppression | VL results at first scheduled VL test | VL result within the 3-month window (56-111 days) or 6-month window (112-251 days)* | Clients in each VL outcome category within the respective 3- or 6-month testing window | Clients with a documented VL result within the respective 3- or 6-month testing window |
| Viral suppression at second scheduled VL test | VL outcomes at second scheduled VL test | VL result within the applicable 10-month (252-307 days) or 12-month (308-391 days) testing window* | Clients in each VL outcome category at the second scheduled VL test. | Clients with a documented second scheduled VL result within the applicable 10- or 12-month testing window |

VL, viral load
\*VL results are categorised as suppressed (<50 copies/mL), low-level viraemia (50–199 copies/mL), intermediate viraemia (200–999 copies/mL), and unsuppressed (≥1,000 copies/mL). Where multiple VL results occurred within a testing window, the result closest to the window's defined milestone (84, 182, 280, and 350 days for the 3-, 6-, 10-, and 12-month windows respectively) was selected.

For all VL endpoints (3 or 6 months and 10 or 12 months), VL was categorised as suppressed (<50 copies/mL), low-level viraemia (50-199 copies/mL), intermediate viraemia (200-999 copies/mL), or unsuppressed (≥1000 copies/mL). To determine whether earlier VL testing at 3 months rather than 6 months was associated with treatment outcomes, we compared the proportion of clients achieving viral suppression at the first and second scheduled VL tests between those initially tested at 3 months and those initially tested at 6 months. We defined the first scheduled VL test as a VL result documented within either the 3-month testing window (56-111 days after ART initiation) or the 6-month testing window (112-251 days after ART initiation) and the second scheduled VL result within the 10-month testing window (252-307 days) or 12-month testing window (308-391 days) after ART initiation.

To assess implementation of the revised six-monthly visit schedule, analyses were restricted to clients who achieved viral suppression (<50 copies/mL) at their first scheduled VL test, as viral suppression was an eligibility criterion for six-monthly clinical visits under the revised guidelines. For these clients, we identified the first clinic visit following the scheduled VL test and calculated the interval between that visit and the next scheduled appointment using recorded appointment dates. Clients with a scheduled appointment <168 days later were classified as having been assigned to <6-month visit intervals. Clients with a scheduled appointment ≥168 days later were classified as having been assigned to ≥6-month visit intervals; those with no recorded clinic visit following their scheduled VL result were classified as having “no visit after VL result.” Clients who disengaged from care were classified using outcome status recorded in TIER.Net (death, loss to follow-up, or transfer to another facility). At each stage of the cascade, clients with no recorded outcome and no subsequent clinic visits after the relevant milestone (first or second scheduled VL test) were classified as having disengaged from care as of their last observed visit.

### Data analysis

We used descriptive statistics to summarise cohort characteristics, reporting frequencies and proportions. Interrupted time-series (ITS) analyses assessed changes in two outcomes following implementation of the revised guidelines in April 2023: uptake of three-month viral load (VL) testing and assignment to extended clinic visit intervals (≥6 months).

The analysis populations differed according to the outcome being assessed. The ITS analysis of three-month VL testing included the eligible ART cohort, whereas the analysis of extended visit-interval assignment was restricted to clients with a suppressed VL result (<50 copies/mL) in the 3-month testing window, reflecting the revised guideline recommendation linking earlier viral suppression to eligibility for extended visit intervals. Descriptive analyses of visit-interval assignment additionally included clients with a suppressed first scheduled VL result in either the 3- or 6-month testing window, allowing comparison of visit-interval assignment by timing of the first VL test.

The unit of analysis for both ITS outcomes was the district-quarter. We modelled three distinct periods: pre-implementation (January 2022–March 2023), implementation (April–December 2023), and post-implementation (January 2024 onward), allowing each period to have its own level and trend. Models were estimated separately by district using segmented linear regression of the quarterly proportion of clients with a VL result in the 3-month testing window and, for the second outcome, the quarterly proportion of virally suppressed clients assigned to extended (≥6-month) visit intervals. Newey-West standard errors with a lag of 1 were used to account for autocorrelation.

The pre-implementation and implementation periods were consistent across both outcomes in King Cetshwayo and Ehlanzeni. In these districts, the post-implementation period comprised six quarters (January 2024-June 2025). In West Rand, where data were available for a shorter period, the post-implementation period comprised five quarters for 3-month VL testing and four quarters for visit-interval assignment.

We compared clients who received their first scheduled VL test during the 3-month window with those who received their first VL test during the 6-month window. We assessed the proportions of clients with documented first and second scheduled VL test results (first at 3 or 6 months and second at 10 or 12 months, respectively), as well as the proportions achieving viral suppression (<50 copies/mL) at each scheduled test.

Among clients retained in care throughout the first year of ART, we summarised the median number of clinic visits and interquartile ranges (IQRs), stratified by calendar year in relation to guideline implementation.

Modified Poisson regression models with robust standard errors were used to estimate relative risks (RRs) and 95% confidence intervals (CIs) for factors associated with VL uptake and viral suppression at the first and second scheduled VL tests. Variables included in adjusted models were selected a priori based on epidemiological relevance, with timing of the first scheduled VL test and assignment to ≥6- month visit intervals considered key exposures of interest. Other demographic and clinical characteristics were included as potential confounders. Clients with missing baseline CD4 count were included as a distinct category in descriptive tables (Table 2) but excluded from regression models via complete-case analysis. All analyses were conducted using Stata version 18 (StataCorp, College Station, TX, USA).

**Table 2.**
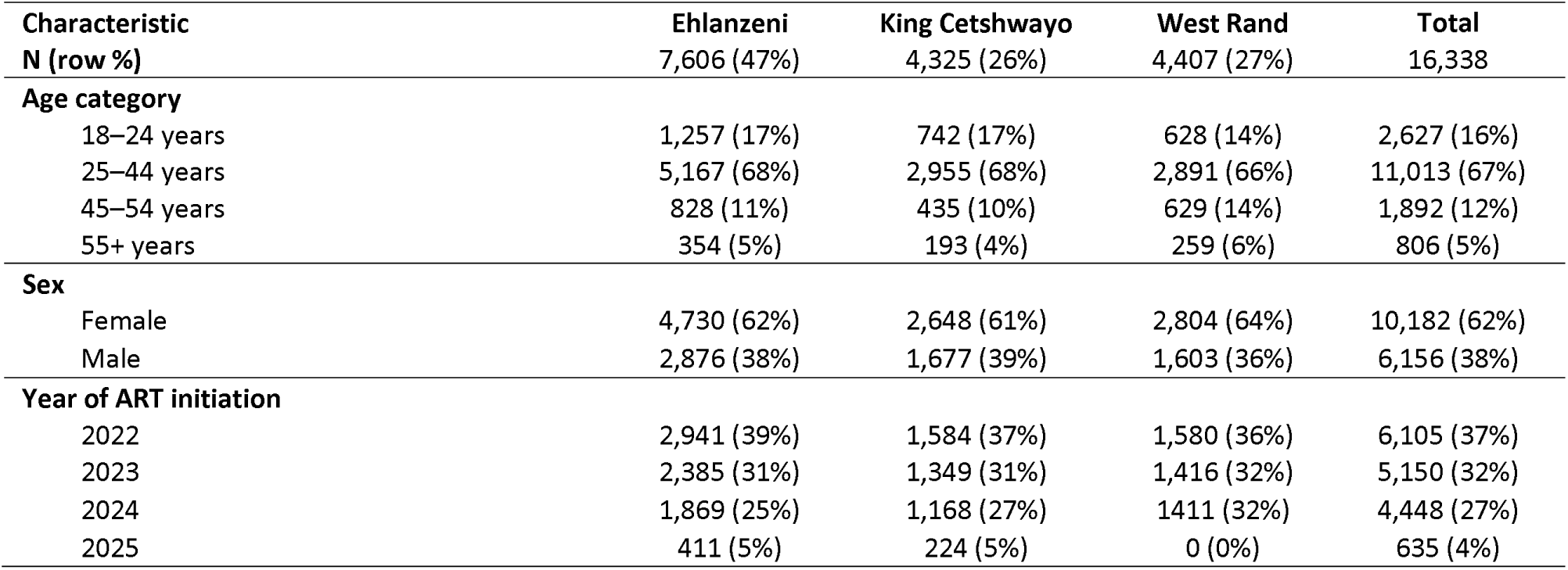
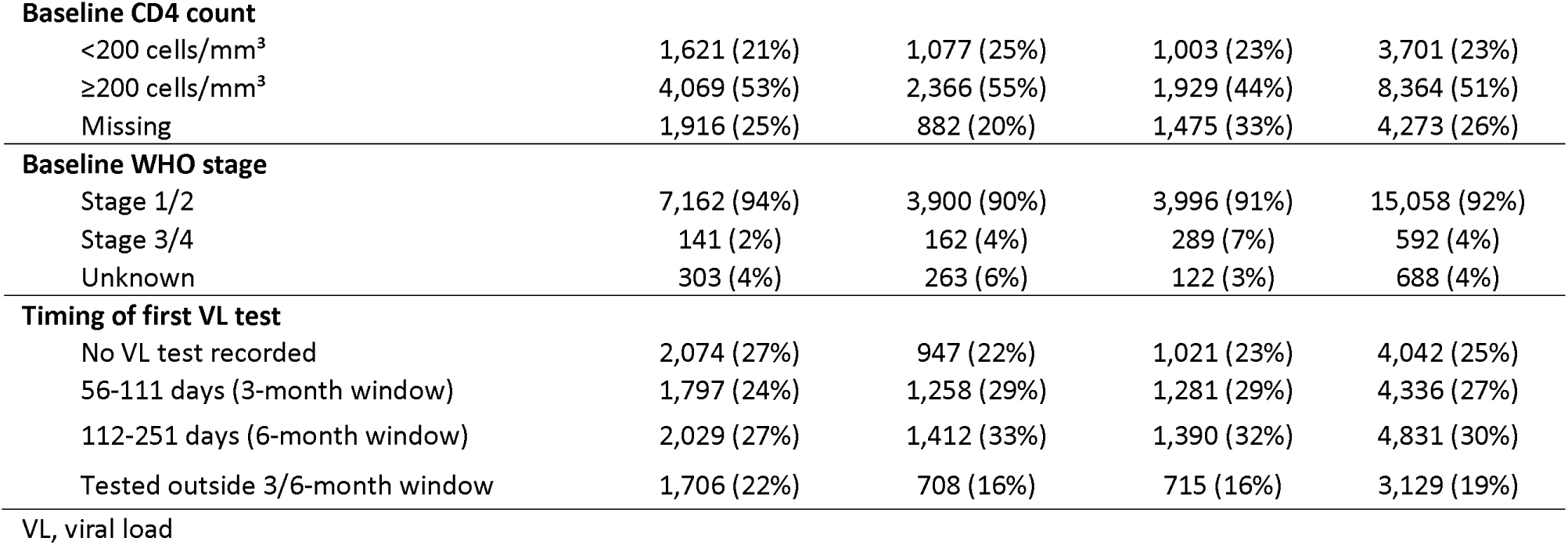
Characteristics of participants initiating or re-initiating ART between January 2022 and March 2025, by district (n=16,338)

| **Characteristic** | **Ehlanzeni** | **King Cetshwayo** | **West Rand** | **Total** |
|---|---|---|---|---|
| **N (row %)** | 7,606 (47%) | 4,325 (26%) | 4,407 (27%) | 16,338 |
| **Age category** |  |  |  |  |
| 18–24 years | 1,257 (17%) | 742 (17%) | 628 (14%) | 2,627 (16%) |
| 25–44 years | 5,167 (68%) | 2,955 (68%) | 2,891 (66%) | 11,013 (67%) |
| 45–54 years | 828 (11%) | 435 (10%) | 629 (14%) | 1,892 (12%) |
| 55+ years | 354 (5%) | 193 (4%) | 259 (6%) | 806 (5%) |
| **Sex** |  |  |  |  |
| Female | 4,730 (62%) | 2,648 (61%) | 2,804 (64%) | 10,182 (62%) |
| Male | 2,876 (38%) | 1,677 (39%) | 1,603 (36%) | 6,156 (38%) |
| **Year of ART initiation** |  |  |  |  |
| 2022 | 2,941 (39%) | 1,584 (37%) | 1,580 (36%) | 6,105 (37%) |
| 2023 | 2,385 (31%) | 1,349 (31%) | 1,416 (32%) | 5,150 (32%) |
| 2024 | 1,869 (25%) | 1,168 (27%) | 1411 (32%) | 4,448 (27%) |
| 2025 | 411 (5%) | 224 (5%) | 0 (0%) | 635 (4%) |
| **Baseline CD4 count** |  |  |  |  |
| <200 cells/mm<sup>3</sup> | 1,621 (21%) | 1,077 (25%) | 1,003 (23%) | 3,701 (23%) |
| ≥200 cells/mm<sup>3</sup> | 4,069 (53%) | 2,366 (55%) | 1,929 (44%) | 8,364 (51%) |
| Missing | 1,916 (25%) | 882 (20%) | 1,475 (33%) | 4,273 (26%) |
| **Baseline WHO stage** |  |  |  |  |
| Stage 1/2 | 7,162 (94%) | 3,900 (90%) | 3,996 (91%) | 15,058 (92%) |
| Stage 3/4 | 141 (2%) | 162 (4%) | 289 (7%) | 592 (4%) |
| Unknown | 303 (4%) | 263 (6%) | 122 (3%) | 688 (4%) |
| **Timing of first VL test** |  |  |  |  |
| No VL test recorded | 2,074 (27%) | 947 (22%) | 1,021 (23%) | 4,042 (25%) |
| 56-111 days (3-month window) | 1,797 (24%) | 1,258 (29%) | 1,281 (29%) | 4,336 (27%) |
| 112-251 days (6-month window) | 2,029 (27%) | 1,412 (33%) | 1,390 (32%) | 4,831 (30%) |
| Tested outside 3/6-month window | 1,706 (22%) | 708 (16%) | 715 (16%) | 3,129 (19%) |

VL, viral load

**Table 3.**
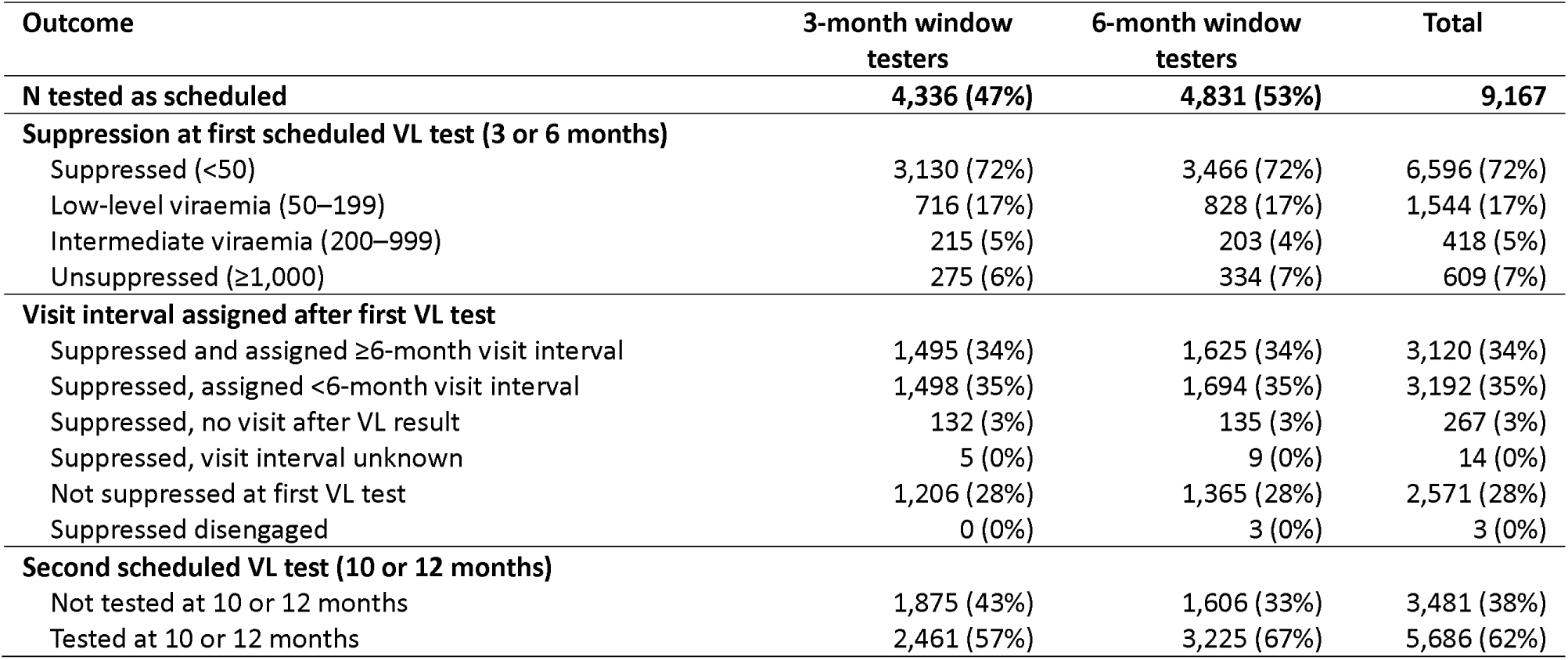
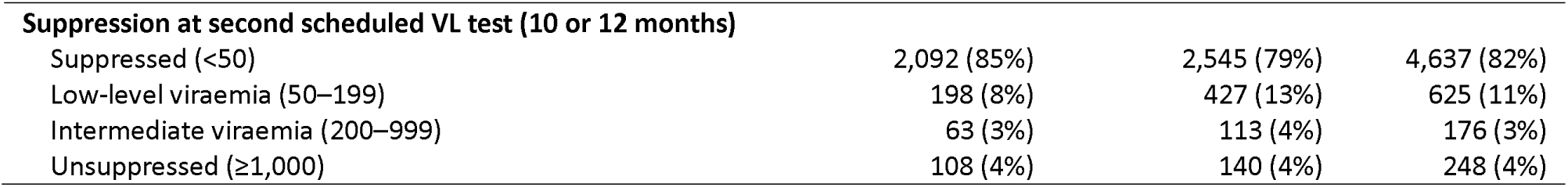
VL test uptake and suppression results among participants tested as scheduled, by timing of tests.

| Outcome | 3-month window testers | 6-month window testers | Total |
|---|---|---|---|
| **N tested as scheduled** | **4,336 (47%)** | **4,831 (53%)** | **9,167** |
| **Suppression at first scheduled VL test (3 or 6 months)** |  |  |  |
| Suppressed (<50) | 3,130 (72%) | 3,466 (72%) | 6,596 (72%) |
| Low-level viraemia (50–199) | 716 (17%) | 828 (17%) | 1,544 (17%) |
| Intermediate viraemia (200–999) | 215 (5%) | 203 (4%) | 418 (5%) |
| Unsuppressed (≥1,000) | 275 (6%) | 334 (7%) | 609 (7%) |
| **Visit interval assigned after first VL test** |  |  |  |
| Suppressed and assigned ≥6-month visit interval | 1,495 (34%) | 1,625 (34%) | 3,120 (34%) |
| Suppressed, assigned <6-month visit interval | 1,498 (35%) | 1,694 (35%) | 3,192 (35%) |
| Suppressed, no visit after VL result | 132 (3%) | 135 (3%) | 267 (3%) |
| Suppressed, visit interval unknown | 5 (0%) | 9 (0%) | 14 (0%) |
| Not suppressed at first VL test | 1,206 (28%) | 1,365 (28%) | 2,571 (28%) |
| Suppressed disengaged | 0 (0%) | 3 (0%) | 3 (0%) |
| **Second scheduled VL test (10 or 12 months)** |  |  |  |
| Not tested at 10 or 12 months | 1,875 (43%) | 1,606 (33%) | 3,481 (38%) |
| Tested at 10 or 12 months | 2,461 (57%) | 3,225 (67%) | 5,686 (62%) |
| **Suppression at second scheduled VL test (10 or 12 months)** |  |  |  |
| Suppressed (<50) | 2,092 (85%) | 2,545 (79%) | 4,637 (82%) |
| Low-level viraemia (50–199) | 198 (8%) | 427 (13%) | 625 (11%) |
| Intermediate viraemia (200–999) | 63 (3%) | 113 (4%) | 176 (3%) |
| Unsuppressed (≥1,000) | 108 (4%) | 140 (4%) | 248 (4%) |

### Artificial intelligence (AI) disclosure statement

ChatGPT 5.6 Sol was utilized during the preparation of this manuscript to design figures from data provided, check internal consistency of data and results reported, edit text for clarity, and copyedit text. The authors collected and analysed all data, reviewed all AI contributions, and confirmed all citations. The authors take full responsibility for the accuracy, integrity, and final content of this publication.

### Ethics approval

The use of de-identified medical record data for this analysis was approved by the Boston University Institutional Review Board (H-38115) and by the University of the Witwatersrand Human Research Ethics Committee (M190445). The protocol was also approved by Provincial Health Research Committees through the National Health Research Database for each study district in South Africa. We received a waiver of informed consent for all medical record data. Authors did not have access to identifying information, as data were provided by the database owner in de-identified format.

## RESULTS

### Characteristics of the study sample

Of the 19,756 clients who initiated or re-initiated ART at the 18 study facilities from January 2022 onward, we excluded 744 who were aged <18 years and 451 who were pregnant. Of the remaining 18,561 clients, 2,223 were excluded because they did not have sufficient individual follow-up time (at least 392 days) to reach the final scheduled VL testing window, based on the end of available data for their district. The final analytic cohort comprised 16,338 clients. (Supplementary Figure 1). As shown in Table 2, 62% of the cohort were female, 67% were aged 25-44 years, and 92% were assigned to WHO stage 1 or 2 at ART initiation. Approximately one-quarter (23%) had a baseline CD4 count <200 cells/mm³, and 26% had no baseline CD4 result recorded.

### Overview of the VL testing and suppression cascade

Figure 3 presents the VL testing and suppression cascade for the full analytic cohort (N=16,338). At the first scheduled VL assessment (Figure 3A), 4,336 (27%) clients had a VL result documented within the 3-month testing window, 4,831 (30%) within the 6-month testing window, 3,129 (19%) were tested outside the scheduled windows, and 4,042 (25%) had no VL result recorded. Overall, 56% of clients had a VL result documented within the recommended 3- or 6-month testing windows. 72% of clients were virally suppressed in both the 3-month testing window (3,130/4,336) and the 6-month window (3,466/4,831).

**Figure 3.**
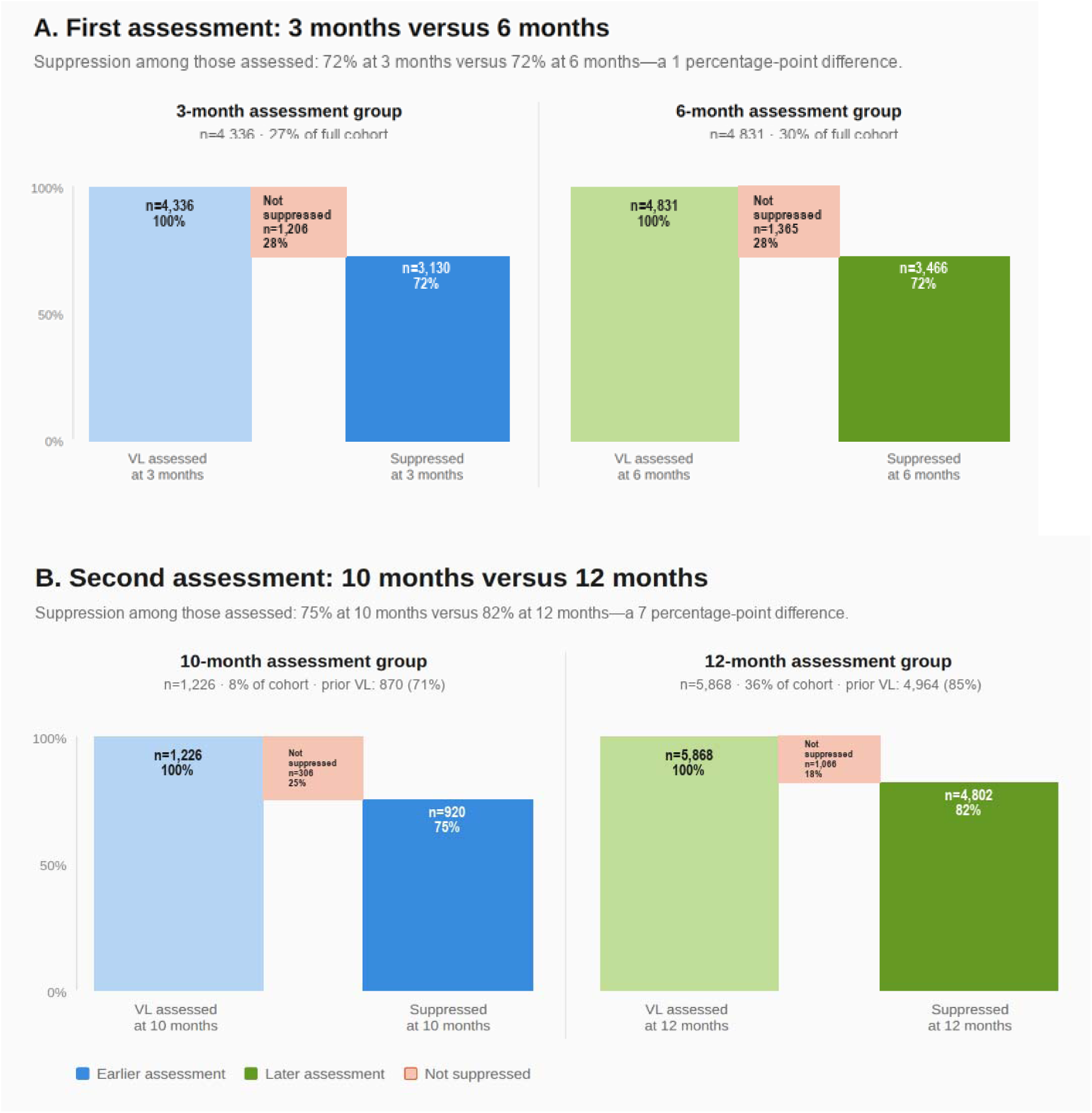
Viral load testing cascade of clients initiated on ART from January 2022 to March 2025.

By the second scheduled assessment at 10 or 12 months (Figure 3B), 6,081/16,338 (37%) clients were no longer in care at the original sites. Of these, 63 had died, 1,257 were lost to follow-up, and 461 had transferred out; the remaining 4,300 were classified as disengaged from care. A further 3,163 (19%) clients remained in care but had no documented second VL result: 338 (2%) had never had a VL test, 1,980 (12%) had a first VL result but no second VL result, and 845 (5%) had been tested only outside the scheduled 3- or 6-month and 10- or 12-month windows. Among the 4,336 clients tested within the 3- month window at the first assessment, 531 (12%) had a subsequent VL result at 10 months and 1,930 (45%) at 12 months. Across the full cohort, 1,226 clients had a VL result at 10 months, of whom 920 (75%) were virally suppressed, while 5,868 had a VL result at 12 months, of whom 4,802 (82%) were virally suppressed (Supplementary Table 3).

### Uptake of first and second scheduled viral load tests

Figure 4 illustrates uptake of 3-month VL testing and 4-month assignment of 6-month visits across the study period. Prior to implementation of the 2023 HIV treatment guidelines, conducting a VL test before six months on ART was relatively unusual; early testing may have reflected individual clinical circumstances or previous ART exposure among clients re-initiating treatment. In all three districts, uptake of early VL testing during the pre-implementation period was below 25% (median 17%).

**Figure 4.**
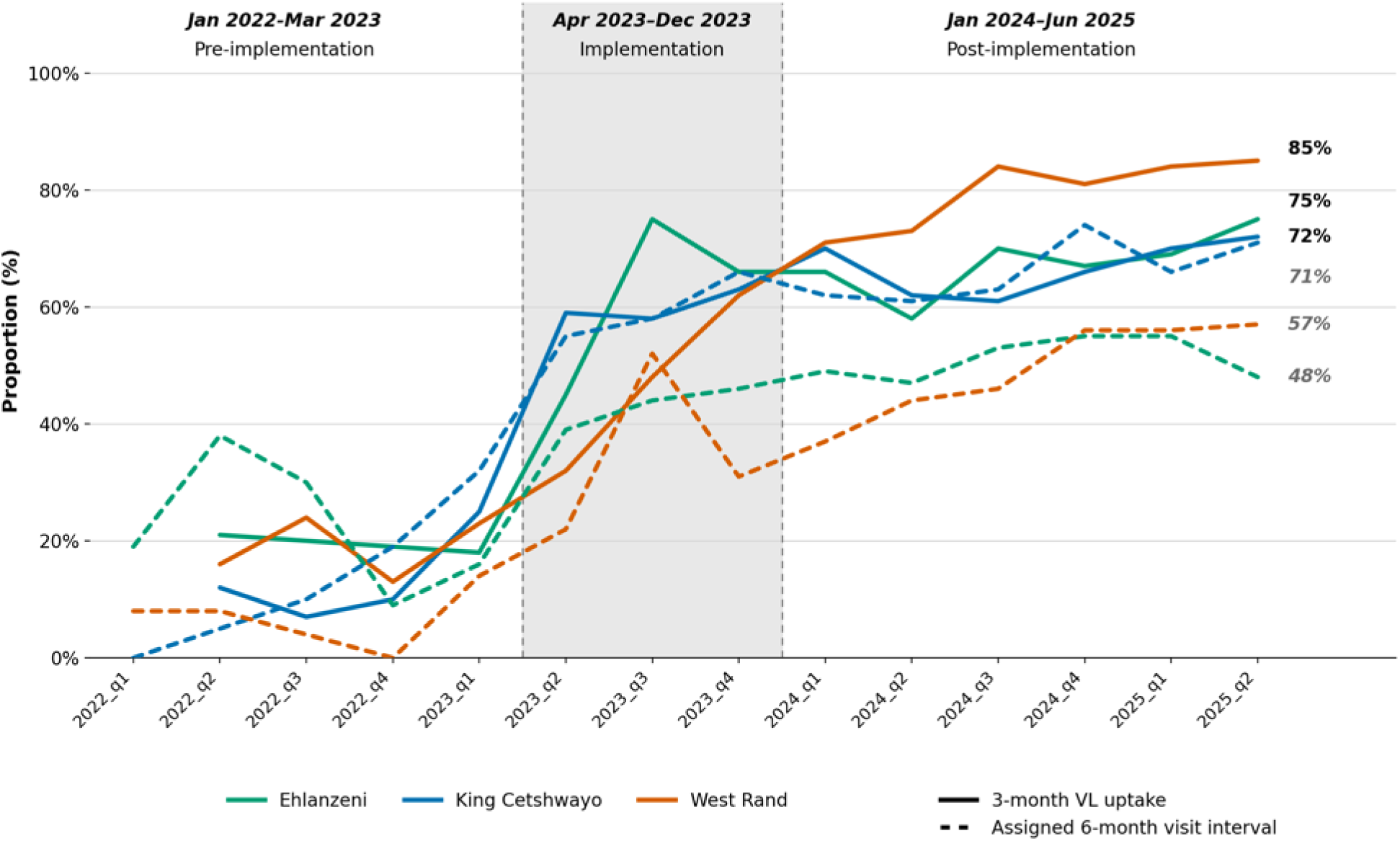
Uptake of three-month viral load test and assignment to ≥6-month visit intervals after suppressed three-month viral load test.

Following guideline implementation in the second quarter of 2023, all districts experienced large, immediate increases in 3-month VL testing uptake, with estimated increases of 43 percentage points in Ehlanzeni (95% CI 26.5–59.1) and King Cetshwayo (34.3–50.8), and 24 percentage points in West Rand (3.3–45.0). Post-implementation trends differed across districts. Coverage continued to increase significantly in West Rand (7% per quarter; 95% CI 3.24–10.71), the steepest sustained trajectory of the three districts, and mildly in King Cetshwayo (1% per quarter; 0.03–1.80), while no significant post-implementation trend was observed in Ehlanzeni (2% per quarter; -0.98 to 4.94), where coverage plateaued after the initial jump.

By the end of the study period, 3-month VL testing coverage had reached 85% in West Rand, 72% in King Cetshwayo, and 75% in Ehlanzeni (Figure 4; Supplementary Table 1).

Among the 6,596 clients who achieved viral suppression at their first VL test, 48% of those tested in the 3-month window and 47% of those tested in the 6-month window were assigned to visit intervals of ≥6 months, while 48% and 49%, respectively, continued with shorter visit intervals; the remainder had no visit after their VL result or an interval that could not be determined (Figure 4).

By the end of the study period in mid-2025, ≥6-month visit intervals were assigned to 57% of virally suppressed clients in West Rand, 71% in King Cetshwayo, and 48% in Ehlanzeni. Post-implementation trends in the three districts varied, with West Rand continuing to expand uptake but little sign of continued increases in the other two districts. (Supplementary Table 2).

Among clients with a first scheduled VL in the 3- or 6-month window (N=9,167), a second scheduled VL test result was documented for 62% overall (5,686/9,167), including 57% of those tested in the 3-month window and of those tested in the 6-month window. In the full cohort (N=16,338), 7,094 (43.4%) clients had a documented VL result within the applicable second scheduled VL testing window, including 1,226 at 10 months and 5,868 at 12 months. This included clients with and without a documented first VL result. Among clients who had a suppressed first scheduled VL within the 3- or 6-month window (N=6,596), uptake of the second scheduled VL was similar among those assigned ≥6-month follow-up intervals and those assigned shorter intervals (RR 1.03, 95% CI 0.97-1.09; aRR 1.02, 95% CI 0.96-1.08).

### Clinic visit frequency during the first year on ART

Between 2022 and 2024, the median number of clinic visits during the first year after ART initiation declined from 8 (IQR: 6-9) in 2022 to 7 (6–8) in 2023 and 6 (5–8) in 2024 across all three districts (Supplementary Figure 4). The distribution of clinic visit frequency shifted progressively towards fewer visits over this period, becoming increasingly concentrated around six to seven visits, with a reduction in the proportion of clients requiring eight or more visits during the first year of ART.

### VL suppression at first and second tests

The distribution of detectable viraemia was comparable, with low-level viraemia (50-199 copies/mL) observed in 16.5% and 17.1% of clients, intermediate viraemia (200-999 copies/mL) in 5.0% and 4.2%, and unsuppressed VL (≥1,000 copies/mL) in 6.3% and 6.9% of clients tested in the 3-month and 6-month windows, respectively.

### Predictors of viral suppression

In both univariate and adjusted modified Poisson regression analyses, viral suppression at the first scheduled VL test was similar among clients tested at 3 months and those tested at 6 months (RR 1.01, 95% CI 0.98–1.03; aRR 1.01, 95% CI 0.98–1.03). Figure 5 illustrates adjusted results, while univariate results are shown in Supplementary Table 4. As is typically reported, male clients had a lower likelihood of viral suppression than female clients (aRR 0.90, 95% CI 0.88-0.93). Viral suppression was higher among clients in King Cetshwayo (1.16, 1.12-1.19) and West Rand (1.11, 1.08-1.15) than in Ehlanzeni, and among clients with a baseline CD4 count ≥200 cells/mm³ compared with those with a baseline CD4 count <200 cells/mm³ (1.23, 1.19-1.27). There were no meaningful differences in viral suppression by age category (RR 0.97–1.00, aRR 1.01–1.04, all close to the null).

**Figure 5.**
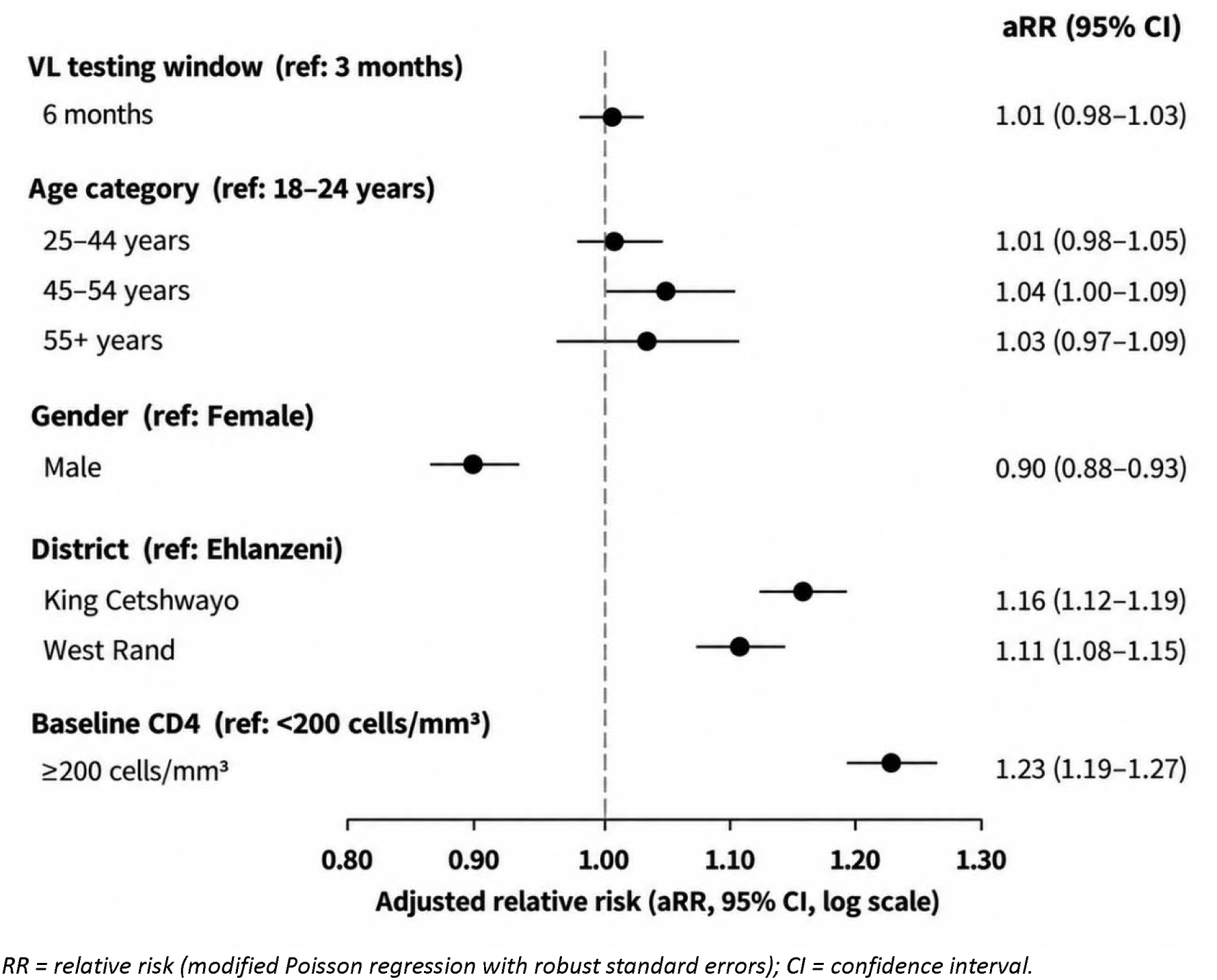
Factors associated with viral suppression (<50 copies/mL) at first scheduled VL test (3- or 6-month), adjusted modified Poisson regression.

At the second scheduled VL test, viral suppression was slightly higher among clients assigned ≥6-month follow-up intervals than among those assigned shorter intervals (91% vs. 89%; RR 1.03, 95% CI 1.00–1.06; aRR 1.02, 95% CI 0.99-1.06) (Figure 6, Supplementary Table 5). Clients with a baseline CD4 count ≥200 cells/mm³ were more likely to achieve viral suppression than those with a baseline CD4 count <200 cells/mm³ (91% vs. 86%; aRR 1.06, 95% CI 1.02-1.11). Viral suppression was also higher among clients receiving care in King Cetshwayo (93% vs. 87%; 1.08, 1.04-1.12) and West Rand (90% vs. 87%; 1.04, 1.00-1.08) than among those in Ehlanzeni. There was little evidence of differences in viral suppression by age category or by sex.

**Figure 6.**
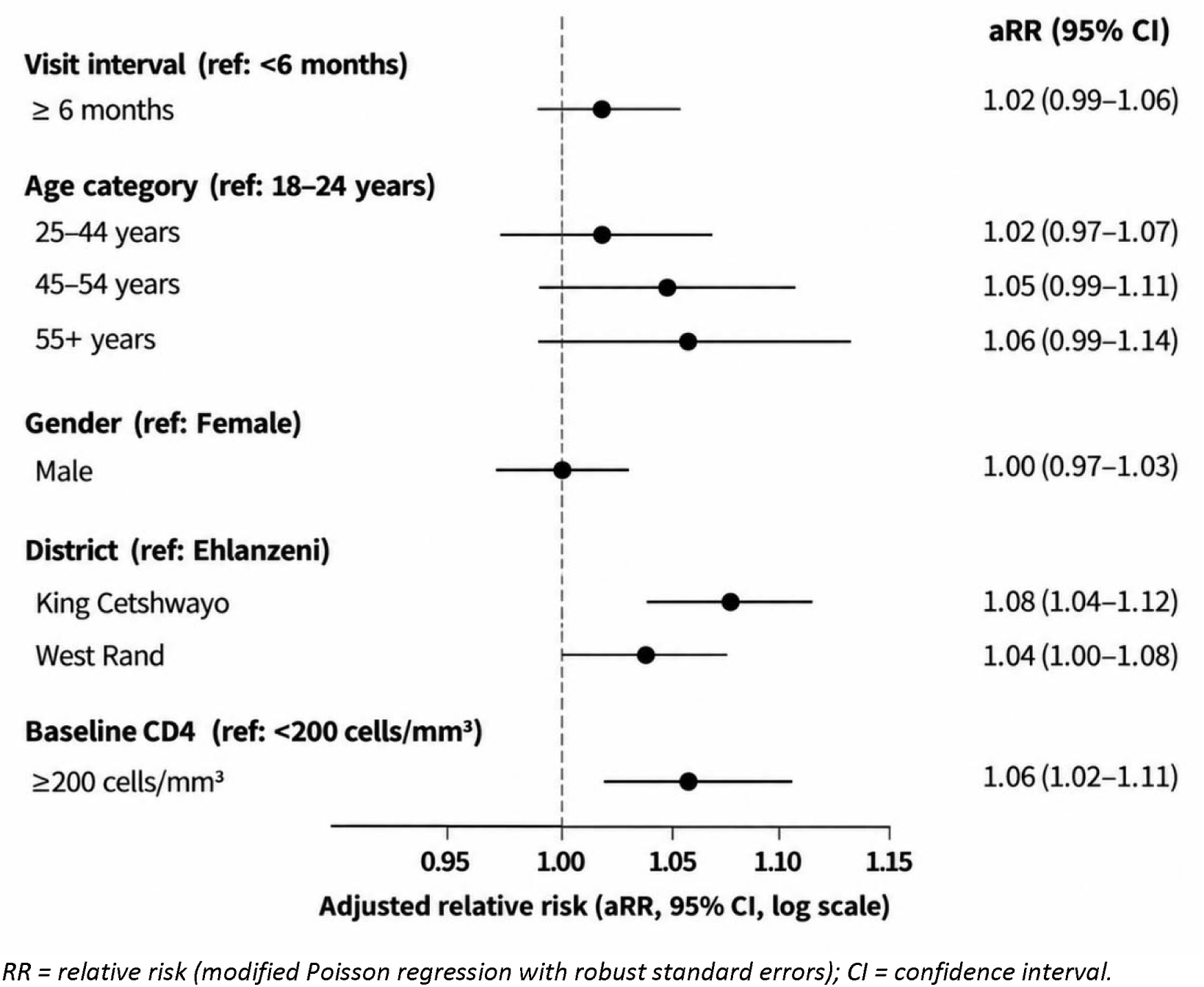
Factors associated with viral suppression at the second scheduled VL test among clients with a documented second VL result.

## DISCUSSION

In this study of 16,338 adult South Africans who initiated ART since January 2022, the introduction of early VL testing and earlier assignment to longer visit intervals was not associated with poorer subsequent virologic outcomes. As intended, the 2023 ART guidelines were associated with rapid uptake of 3-month VL testing, earlier assignment to longer visit intervals, and fewer clinic visits during the first year on treatment. Together, these findings suggest that earlier de-intensification of care can be implemented without sacrificing viral suppression among clients who achieve early viral suppression.

South Africa’s experience demonstrates that early (three-month) VL testing is operationally feasible and can be implemented at scale within routine primary healthcare settings. South Africa is not the only country to have moved towards earlier monitoring: Kenya’s national guidelines have recommended 3-month VL testing for ART initiators since 2022(21). Under the Kenyan schedule, adult clients (>25 years) are tested at 3 and 12 months and annually thereafter for those suppressed. Notably, however, Kenya’s DSD eligibility criteria still require six months on treatment, separating earlier VL monitoring from early assignment to less intensive care. Our finding that earlier VL testing combined with early assignment to longer prescribing and visit intervals was not associated with poorer subsequent outcomes supports the feasibility of allowing clients earlier access to differentiated care. These findings should be interpreted with caution, however, as the observed changes followed guideline implementation but cannot be attributed to the guidelines alone. In the absence of a comparison group, concurrent changes in service delivery or other factors may also have contributed to the observed trends.

Early assignment to longer visit intervals has previously been used pragmatically elsewhere in sub-Saharan Africa during periods of disruption, including the 2014–2016 Ebola outbreak, armed conflict, and the COVID-19 pandemic, to maintain continuity of treatment while reducing facility contact(23). Our findings provide evidence that early assignment to longer visit intervals can also be implemented in routine, non-emergency settings and that the historical requirement for clients to be on ART for 6 or 12 months before becoming eligible for DSD models(22) may not be necessary for clients who have achieved early viral suppression.

The reduction in clinic visits observed in our cohort, from a median of 8 first-year visits in 2022 to 6 in 2024, should reduce the burden associated with engaging in HIV care and is also consistent with client preferences: in a discrete choice experiment conducted among clients at our study sites, clients expressed a preference for longer dispensing intervals(24). We could not directly assess whether the reduction in visit frequency itself was associated with subsequent VL monitoring or viral suppression. Frequent clinic attendance and the associated travel costs, opportunity costs, clinic waiting times, and consequences of missed appointments are well-documented barriers to sustained engagement in HIV care(25,26). Clients have reported that care-seeking can be particularly burdensome during the early treatment period, when visit frequency has historically been highest(27).

This study had several limitations. First, approximately one quarter of clients had no documented VL test. We cannot determine whether this reflects incomplete data capture in TIER.Net or incomplete VL testing, and our estimates of guideline fidelity may be biased if tests were performed according to guideline but were not captured in our data. If missingness is due primarily to incomplete data capture-that is, tests were performed but not recorded-overall suppression could be higher than observed among clients with documented results. If missingness is due largely to incomplete testing, results may instead overstate population-level suppression if failure to obtain a VL test is associated with treatment interruption or disengagement. Furthermore, estimates of viral suppression were restricted to clients with documented VL results and may therefore not represent outcomes among all eligible clients. If the likelihood of having a documented result is associated with retention in care, adherence, or clinical status, the observed suppression estimates may be subject to selection bias.

Second, because the TIER.Net DMOC enrolment field was incompletely captured, we relied on assignment to ≥6-month visit intervals as proxy measures of differentiated service delivery uptake. These proxies may overestimate formal DMOC enrolment because clients can receive extended visit or dispensing intervals without enrolment in a DMOC. Third, the study was conducted at 18 facilities in three districts, and, as such, our results may not be generalizable to other South African facilities and locations with different levels of implementation support, staffing, or infrastructure. Fourth, although we adjusted the comparisons of visit interval and VL-testing timing for observed demographic and clinical characteristics, residual confounding by important unmeasured variables remains possible. Clients assigned longer visit intervals may differ systematically from those assigned shorter intervals in characteristics not captured in routine data, including adherence history, clinical stability, or clinician assessment of readiness for differentiated care. Fifth, analyses of viral suppression at the second scheduled VL test were necessarily restricted to clients with a documented second VL result. Although availability of the second scheduled VL did not differ substantially by initial VL test timing, selection related to VL ascertainment cannot be excluded. Finally, we were unable to distinguish reliably between ART-naïve initiators and clients re-initiating treatment and therefore could not assess whether the associations observed differed by prior ART exposure. This may be particularly relevant to the pre-implementation period, when earlier VL testing may have been more common among clients re-initiating treatment.

## CONCLUSION

South Africa’s revised 2023 ART guidelines were associated with rapid uptake of earlier VL monitoring, earlier assignment to longer visit intervals, and fewer clinic visits during the first year of ART, although implementation remained incomplete. Among clients with documented follow-up VL results, assignment to longer visit intervals was not associated with poorer subsequent viral suppression. Continued efforts to improve implementation and VL documentation are warranted, alongside evaluation of longer-term retention and treatment outcomes.

## Author contributions

Conceptualization: EK, NS, AM, MM, SR

Data curation: VN, EK

Formal analysis: EK

Funding acquisition: SR

Methodology: EK NS MB AM MM SR

Project administration: LS, VN

Supervision: SR LS

Writing – original draft: EK

Writing – review & editing: LS, NM, VN, MB, LM, SR, MM, NS, AM

## Competing interests

The authors have declared that no competing interests exist. LM and MM are employees of the government agency that has supervisory authority over the study sites.

## Funding

Funding for the study was provided by the Gates Foundation through award INV-031690 to Boston University. The funder had no role in study design, data collection, analysis, or preparation of this manuscript.

## Data availability

TIER.Net data are owned by the South African National Department of Health. They can be requested from Mr. Mbulelo Cabuko, Director, Health Information Research Monitoring and Evaluation (HIRME) Cluster, National Department of Health, Pretoria, South Africa. Analytic code for this analysis will be shared on GitHub once the manuscript is published.

## Supplementary files

### Supplementary figures

1. Supplementary Figure 1. Study sample flow chart.
2. Supplementary Figure 2. Factors associated with uptake of the second scheduled VL test (10 or 12 months) among all clients with a documented result, regardless of first-VL testing window, adjusted and unadjusted modified Poisson regression.
3. Supplementary Figure 3. Factors associated with viral suppression (<50 copies/mL) at 10 or 12 months among all clients with a documented result, regardless of first-VL testing window, adjusted and unadjusted modified Poisson regression.
4. Supplementary Figure 4. Number of clinic visits in first year on ART, year of initiation, 2022– 2024.

### Supplementary tables

1. Supplementary Table 1. Interrupted time series results for 3-month VL testing coverage, by district.
2. Supplementary Table 2. Interrupted time series results for uptake of ≥6-month visit intervals, by district.
3. Supplementary Table 3. VL testing and suppression cascade for the full analytic cohort (N=16,338), by timing of first scheduled VL test.
4. Supplementary Table 4. Unadjusted (univariable) associations with VL suppression at first scheduled VL test.
5. Supplementary Table 5>. Unadjusted (univariable) associations with VL suppression at second scheduled VL test.

## References

1. World Health Organization. Information sheet: HIV statistics, globally and by WHO region, 2025 [Internet]. 2025 [cited 2026 Apr 8]. Available from: https://www.who.int/teams/global-hiv-hepatitis-and-stis-programmes/hiv/strategic-information/hiv-data-and-statistics

2. World Health Organization. Supporting re-engagement in HIV treatment services: Policy brief.

3. Zheng A, Kileel EM, Brennan AT, Flynn DB, Rosen S, Fox MP, et al. Systematic review and meta-analysis of retention and disengagement after initiation on antiretroviral therapy in low-and middle-income countries after the introduction of Universal Test and Treat policies. J Int AIDS Soc. 2025;28:70026. doi:10.1002/jia2.70026/full

4. Nicol E, Basera W, Mukumbang FC, Cheyip M, Mthethwa S, Lombard C, et al. Linkage to HIV Care and Early Retention in Care Rates in the Universal Test-and-Treat Era: A Population-based Prospective Study in KwaZulu-Natal, South Africa. AIDS Behav. 2023 Apr 1;27(4):1068–81. doi:10.1007/s10461-022-03844-w PubMed PMID: 36098845.

5. Maskew M, Benade M, Huber A, Pascoe S, Sande L, Malala L, et al. Patterns of engagement in care during clients’ first 12 months after HIV treatment initiation in South Africa: A retrospective cohort analysis using routinely collected data. PLOS Global Public Health. 2024 Feb 28;4(2):e0002956. doi:10.1371/journal.pgph.0002956

6. Benade M, Maskew M, Chilembo P, Mwansa MW, Savory T, Nichols B, et al. Patterns of engagement in care during clients’ first 12 months after HIV treatment initiation in Zambia: a retrospective cohort analysis using routinely collected data. BMJ Glob Health. 2025;10:17930. doi:10.1136/bmjgh-2024-017930

7. Tweya H, Thawani A, Klabbers R, Huwa J, Rambiki E, Viola E, et al. Tracing the journey: Patient engagement in long-term antiretroviral therapy care in Malawi. PLoS One. 2026 Jun 1;21(6 June). doi:10.1371/journal.pone.0323983 PubMed PMID: 42224296.

8. Lilian RR, Rees K, McIntyre JA, Struthers HE, Peters RPH. Same-day antiretroviral therapy initiation for HIV-infected adults in South Africa: Analysis of routine data. PLoS One. 2020 Jan 1;15(1):e0227572. doi:10.1371/JOURNAL.PONE.0227572 PubMed PMID: 31935240.

9. Hoffman RM, Moyo C, Balakasi KT, Siwale Z, Hubbard J, Bardon A, et al. Multimonth dispensing of up to 6 months of antiretroviral therapy in Malawi and Zambia (INTERVAL): a cluster-randomised, non-blinded, non-inferiority trial. Lancet Glob Health. 2021 May 1;9(5):e628–38. doi:10.1016/S2214-109X(21)00039-5 PubMed PMID: 33865471.

10. Labhardt ND, Brown JA, Sass N, Ford N, Rosen S. Treatment Outcomes after Offering Same-Day Initiation of Human Immunodeficiency Virus Treatment-How to Interpret Discrepancies between Different Studies. Clinical Infectious Diseases. Oxford University Press; 2023. p. 1176–84. doi:10.1093/cid/ciad317 PubMed PMID: 37229594.

11. Huber AN, Jamieson L, Fox MP, Manganye M, Malala L, Chidarikire T, et al. Evaluating the impact of differentiated service delivery (DSD) on retention in care and HIV viral suppression in South Africa: A target trial emulation using routine healthcare data. Geng EH, editor. PLoS Med. 2025 Aug 26;22(8):e1004489. doi:10.1371/journal.pmed.1004489

12. MINIMUM DIFFERENTIATED MODELS OF CARE PACKAGE TO SUPPORT LINKAGE TO CARE, ADHERENCE AND RETENTION IN CARE DIFFERENTIATED MODELS OF CARE STANDARD OPERATING PROCEDURES Adherence Guidelines for HIV, TB and NCDs.

13. Rosen S, Grimsrud A, Ehrenkranz P, Katz I. Models of service delivery for optimizing a patient’s first six months on antiretroviral therapy for HIV: An applied research agenda. Gates Open Res. 2020;4:1–15. doi:10.12688/gatesopenres.13159.1

14. Pascoe S, Huber A, Mokhele I, Lekodeba N, Ntjikelane V, Sande L, et al. The SENTINEL study of differentiated service delivery models for HIV treatment in Malawi, South Africa, and Zambia: research protocol for a prospective cohort study. BMC Health Serv Res. 2023 Aug 23;23(1):891. doi:10.1186/s12913-023-09813-w PubMed PMID: 37612720.

15. Maskew M, Ntjikelane V, Juntunen A, Scott N, Benade M, Sande L, et al. Preferences for services in a patient’s first six months on antiretroviral therapy for HIV in South Africa and Zambia (PREFER): research protocol for a prospective observational cohort study. Gates Open Res. 2023 Sep 12;7:119. doi:10.12688/gatesopenres.14682.1

16. Mokhele I, Ntjikelane V, Scott N, Kaiser J, Morgan A, Huber A, et al. Are HIV Treatment Clients Offered a Choice of Differentiated Service Delivery Models? Evidence from Malawi, South Africa, and Zambia. Patient Prefer Adherence. 2025 Jun;Volume 19:1825–41. doi:10.2147/PPA.S494679

17. Ntjikelane V, Huber A, Morgan A, Pascoe S, Manganye M, Malala L, et al. Primary healthcare providers’ perspectives on six-month dispensing of antiretroviral therapy (ART) in South Africa: cross-sectional survey of views and preferences. Glob Health Action. 2025 Dec 31;18(1). doi:10.1080/16549716.2025.2484111

18. Mutanda N, Morgan A, Kamanga A, Sande L, Ntjikelane V, Maskew M, et al. Experiences and Preferences in Zambia and South Africa for Delivery of HIV Treatment During a Client’s First Six Months: Results of the PREFER Study’s Cross-Sectional Baseline Survey. AIDS Behav. 2025 Jun 1;29(6):1713–28. doi:10.1007/s10461-025-04640-y

19. Republic of South Africa National Department of Health. 2023 ART Clinical Guidelines for the Management of HIV in Adults, Pregnancy and Breastfeeding, Adolescents, Children, Infants and Neonates [Internet]. Pretoria; 2023 Jun [cited 2025 May 19]. Available from: https://knowledgehub.health.gov.za/system/files/elibdownloads/2023-07/National%20ART%20Clinical%20Guideline%20AR%204.5%2020230713%20Version%204%20WEB.pdf

20. 2023 ART Clinical Guidelines for the Management of HIV in Adults, Pregnancy and Breastfeeding, Adolescents, Children, Infants and Neonates | Department of Health Knowledge Hub [Internet]. [cited 2025 Feb 13]. Available from: https://knowledgehub.health.gov.za/elibrary/2023-art-clinical-guidelines-management-hiv-adults-pregnancy-and-breastfeeding-adolescents

21. Kenya HIV Prevention and Treatment Guidelines, 2022 2022 Edition.

22. Larson BA, Pascoe SJ, Huber A, Long LC, Murphy J, Miot J, et al. Will differentiated care for stable HIV patients reduce healthcare systems costs? [Internet]. 2020. doi:10.1002/jia2.25541/full

23. Wilkinson L, Grimsrud A. The time is now: expedited HIV differentiated service delivery during the COVID-19 pandemic. J Int AIDS Soc. 2020 May 1;23(5):e25503. doi:10.1002/JIA2.25503 PubMed PMID: 32378345.

24. Morgan A, Sande L, Maskew M, Rosen S, Long L, Mutanda N, et al. Client preferences for service delivery during the early treatment period in South Africa and Zambia: Mixed-method findings from a discrete choice experiment and concurrent focus group discussions [Internet]. 2025. Available from: http://medrxiv.org/lookup/doi/10.1101/2025.07.28.25332159 doi:10.1101/2025.07.28.25332159

25. Magura J, Nhari SR, Nzimakwe TI. Barriers to ART adherence in sub-Saharan Africa: a scoping review toward achieving UNAIDS 95-95-95 targets. Front Public Health. 2025 Jun 10;13:1609743. doi:10.3389/FPUBH.2025.1609743/TEXT PubMed PMID: 40556913.

26. Hoffmann CJ, Milovanovic M, Kinghorn A, Kim HY, Motlhaoleng K, Martinson NA, et al. Value stream mapping to characterize value and waste associated with accessing HIV care in South Africa. PLoS One. 2018 Jul 1;13(7):e0201032. doi:10.1371/JOURNAL.PONE.0201032 PubMed PMID: 30040836.

27. Maskew M, Mutanda N, Scott N, Morgan A, Benade M, Ntjikelane V, et al. Drivers of disengagement from care during the first six months on antiretroviral therapy for HIV in South Africa: A mixed-methods study. PLOS Global Public Health. 2025 Dec 1;5(12):e0004977. doi:10.1371/JOURNAL.PGPH.0004977

