## Supplementary files for "Uptake and outcomes of earlier viral load testing and longer visit intervals in the early HIV treatment period: retrospective cohort analysis of South Africa’s 2023 ART guidelines revision"

**Supplementary tables**

Supplementary Table 1. Interrupted time series results for 3-month VL testing coverage, by district.

Supplementary Table 2. Interrupted time series results for uptake of ≥6-month visit intervals, by district.

Supplementary Table 3. VL testing and suppression cascade for the full analytic cohort (N=16,338), by timing of first scheduled VL test.

Supplementary Table 4. Unadjusted (univariable) associations with VL suppression at first scheduled VL test.

Supplementary Table 5. Unadjusted (univariable) associations with VL suppression at second scheduled VL test.

**Supplementary figures**

Supplementary Figure 1. Study sample flow chart.

Supplementary Figure 2. Factors associated with uptake of the second scheduled VL test (10 or 12 months) among all clients with a documented result, regardless of first-VL testing window, adjusted and unadjusted modified Poisson regression.

Supplementary Figure 3. Factors associated with viral suppression (<50 copies/mL) at 10 or 12 months among all clients with a documented result, regardless of first-VL testing window, adjusted and unadjusted modified Poisson regression.

Supplementary Figure 4. Number of clinic visits in first year on ART, year of initiation, 2022–2024.

**Supplementary Table 1. Interrupted time series results for 3-month VL testing coverage, by district**

| **District** | **Pre-implementation trend, pp/quarter (95% CI)** | **Percentage increase at implementation, pp (95% CI)** | **Coverage at end of study period** |
| --- | --- | --- | --- |
| **Ehlanzeni** | −1.37 (−3.53 to 0.79) | 42.8 (26.5 to 59.1) † | 72% |
| **King Cetshwayo** | 2.12 (0.28 to 3.97) † | 42.6 (34.3 to 50.8) † | 75% |
| **West Rand** | −0.18 (−3.04 to 2.68) | 24.1 (3.3 to 45.0) † | 85% |

*pp = percentage points; CI = confidence interval.*

*† 95% CI excludes 0*

**Supplementary Table 2. Interrupted time series results for uptake of ≥6-month visit intervals, by district**

| **District** | **Pre-implementation trend, pp/quarter (95% CI)** | **Percentage increase at implementation, pp (95% CI)** | **Coverage at end of study period** |
| --- | --- | --- | --- |
| **Ehlanzeni** | −2.4 (−11.3 to 6.6) | 39.0 (8.1 to 69.8) † | 48% |
| **King Cetshwayo** | 8.5 (3.9 to 13.2) † | 34.1 (7.1 to 61.1) † | 71% |
| **West Rand** | 0.5 (−3.8 to 4.9) | 31.6 (−2.4 to 65.6) | 57% |

*pp = percentage points; CI = confidence interval.*

**Supplementary Table 3. VL testing and suppression cascade for the full analytic cohort (N=16,338), by timing of first scheduled VL test**

| **Outcome** | **No VL test recorded (N=4,042)** | **56-<112 days (3-month window) (N=4,336)** | **112-<252 days (6-month window) (N=4,831)** | **Tested outside window (N=3,129)** | **Total (N=16,338)** |
| --- | --- | --- | --- | --- | --- |
| **VL suppression at first scheduled VL test** | | | | | |
| Suppressed at 3 months | 0 (0.0%) | 3130 (72.2%) | 0 (0.0%) | 0 (0.0%) | 3130 (19.2%) |
| Suppressed at 6 months | 0 (0.0%) | 0 (0.0%) | 3466 (71.7%) | 0 (0.0%) | 3466 (21.2%) |
| Not suppressed at 3 months | 0 (0.0%) | 1206 (27.8%) | 0 (0.0%) | 0 (0.0%) | 1206 (7.4%) |
| Not suppressed at 6 months | 0 (0.0%) | 0 (0.0%) | 1365 (28.3%) | 0 (0.0%) | 1365 (8.4%) |
| No VL, in care | 338 (8.4%) | 0 (0.0%) | 0 (0.0%) | 2257 (72.1%) | 2595 (15.9%) |
| Disengaged without VL | 3704 (91.6%) | 0 (0.0%) | 0 (0.0%) | 872 (27.9%) | 4576 (28.0%) |
| **Visit interval after suppressed first VL** | | | | | |
| ≥6mo among suppressed | 0 (0.0%) | 1495 (34.5%) | 1625 (33.6%) | 0 (0.0%) | 3120 (19.1%) |
| <6mo among suppressed | 0 (0.0%) | 1498 (34.5%) | 1694 (35.0%) | 0 (0.0%) | 3192 (19.5%) |
| Not suppressed at first VL test | 0 (0.0%) | 1206 (27.8%) | 1365 (28.3%) | 0 (0.0%) | 2571 (15.7%) |
| No VL done, still in care | 338 (8.4%) | 0 (0.0%) | 0 (0.0%) | 2257 (72.1%) | 2595 (15.9%) |
| No visits after VL | 0 (0.0%) | 132 (3.0%) | 135 (2.8%) | 0 (0.0%) | 267 (1.6%) |
| Disengaged | 3704 (91.6%) | 0 (0.0%) | 3 (0.1%) | 872 (27.9%) | 4579 (28.0%) |
| VL done, visit interval unknown (no next appointment recorded) | 0 (0.0%) | 5 (0.1%) | 9 (0.2%) | 0 (0.0%) | 14 (0.1%) |
| **Second scheduled VL test** | | | | | |
| Tested at 10m | 0 (0.0%) | 531 (12.2%) | 309 (6.4%) | 30 (1.0%) | 870 (5.3%) |
| Tested at 12m | 0 (0.0%) | 1928 (44.5%) | 2914 (60.3%) | 122 (3.9%) | 4964 (30.4%) |
| No prior VL but tested at 10m | 0 (0.0%) | 0 (0.0%) | 0 (0.0%) | 356 (11.4%) | 356 (2.2%) |
| No prior VL but tested at 12m | 0 (0.0%) | 0 (0.0%) | 0 (0.0%) | 904 (28.9%) | 904 (5.5%) |
| Never had any VL, still in care | 338 (8.4%) | 0 (0.0%) | 0 (0.0%) | 0 (0.0%) | 338 (2.1%) |
| Had 3/6mo VL, no second VL, still in care | 0 (0.0%) | 1137 (26.2%) | 843 (17.4%) | 0 (0.0%) | 1980 (12.1%) |
| Tested outside 3/6mo window only, still in care | 0 (0.0%) | 0 (0.0%) | 0 (0.0%) | 845 (27.0%) | 845 (5.2%) |
| Disengaged | 3704 (91.6%) | 740 (17.1%) | 765 (15.8%) | 872 (27.9%) | 6081 (37.2%) |
| **Suppression at second scheduled VL test** | | | | | |
| Suppressed at 10 months | 0 (0.0%) | 440 (10.1%) | 203 (4.2%) | 277 (8.9%) | 920 (5.6%) |
| Suppressed at 12 months | 0 (0.0%) | 1650 (38.1%) | 2340 (48.4%) | 812 (26.0%) | 4802 (29.4%) |
| Not suppressed at 10 months | 0 (0.0%) | 91 (2.1%) | 106 (2.2%) | 109 (3.5%) | 306 (1.9%) |
| Not suppressed at 12 months | 0 (0.0%) | 278 (6.4%) | 574 (11.9%) | 214 (6.8%) | 1066 (6.5%) |
| Never had any VL, still in care | 338 (8.4%) | 0 (0.0%) | 0 (0.0%) | 0 (0.0%) | 338 (2.1%) |
| Had 3/6mo VL, no second VL, still in care | 0 (0.0%) | 1137 (26.2%) | 843 (17.4%) | 0 (0.0%) | 1980 (12.1%) |
| Tested outside 3/6mo window only, still in care | 0 (0.0%) | 0 (0.0%) | 0 (0.0%) | 845 (27.0%) | 845 (5.2%) |
| Disengaged without VL | 3704 (91.6%) | 740 (17.1%) | 765 (15.8%) | 872 (27.9%) | 6081 (37.2%) |

### Supplementary Table 4. Unadjusted (univariate) associations with VL suppression at first test— companion to Figure 5, which shows adjusted estimates only

| **Characteristic** | **Level** | **Univariate RR** | **95% CI** |
| --- | --- | --- | --- |
| VL testing window | 3 months (ref) | - | - |
|  | 6 months | 1.01 | (0.98–1.03) |
| Age category | 18–24 years (ref) | - | - |
|  | 25–44 years | 0.97 | (0.93–1.00) |
|  | 45–54 years | 0.98 | (0.94–1.02) |
|  | 55+ years | 0.97 | (0.91–1.03) |
| Gender | Female (ref) | - | - |
|  | Male | 0.88 | (0.86–0.91) |
| District | Ehlanzeni (ref) | - | - |
|  | King Cetshwayo | 1.15 | (1.12–1.19) |
|  | West Rand | 1.10 | (1.07–1.14) |
| Baseline CD4 | <200 cells/mm³ (ref) | - | - |
|  | ≥200 cells/mm³ | 1.24 | (1.20–1.28) |

### Supplementary Table 5. Unadjusted (univariate) associations with VL suppression at second test companion to Figure 6, which shows adjusted estimates only

| **Characteristic** | **Level** | **Univariate RR** | **95% CI** |
| --- | --- | --- | --- |
| Visit interval | <6 months (ref) | - | - |
|  | ≥6 months | 1.03 | 1.00–1.06 |
| Age category | 18–24 years (ref) | - | - |
|  | 25–44 years | 1.00 | 0.96–1.04 |
|  | 45–54 years | 1.02 | 0.97–1.08 |
|  | 55+ years | 1.04 | 0.97–1.11 |
| Gender | Female (ref) | - | - |
|  | Male | 1.00 | 0.97–1.03 |
| District | Ehlanzeni (ref) | - | - |
|  | King Cetshwayo | 1.08 | 1.04–1.12 |
|  | West Rand | 1.04 | 1.00–1.08 |
| Baseline CD4 | <200 cells/mm³ (ref) | - | - |
|  | ≥200 cells/mm³ | 1.06 | 1.02–1.11 |

**Supplementary Figure 1. Study sample flow chart**

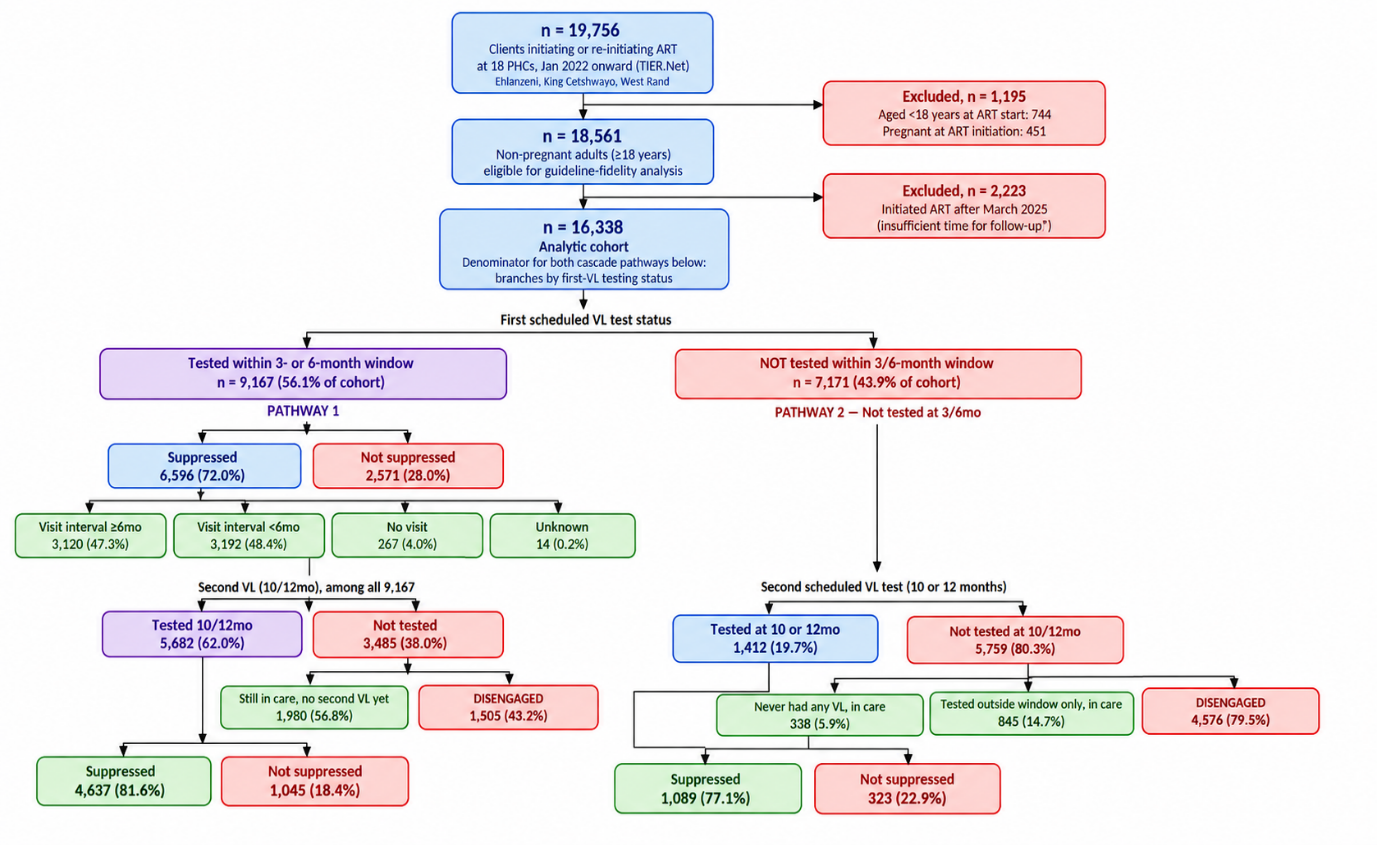

**Supplementary Figure 2. Factors associated with uptake of the second scheduled VL test (10 or 12 months) among all eligible clients, regardless of first VL testing, adjusted and unadjusted modified Poisson regression.**
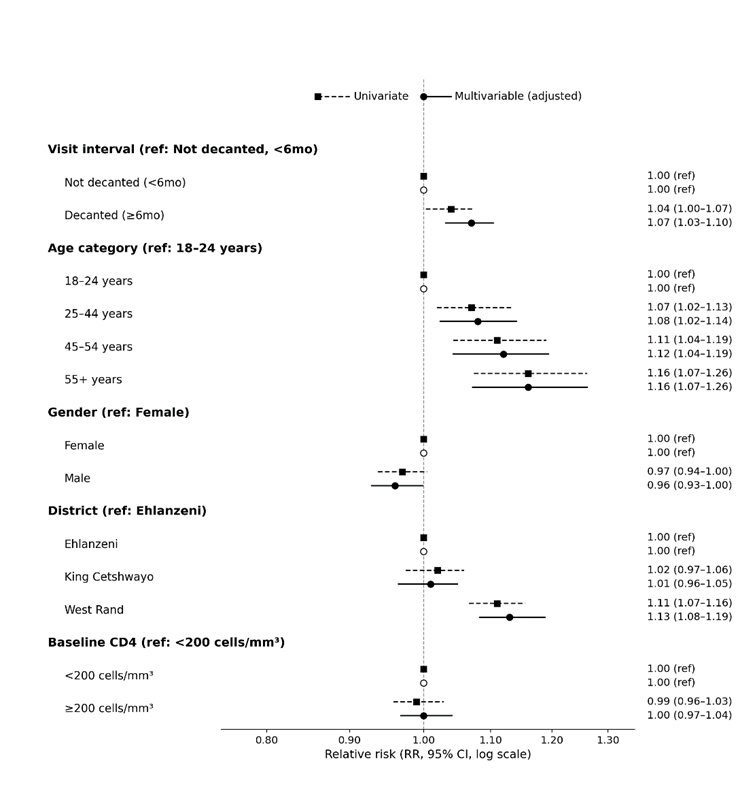

### Supplementary Figure 3. Factors associated with viral suppression (<50 copies/mL) at the second scheduled VL test (10 or 12 months) among all clients with a documented second VL result, regardless of first-VL testing, adjusted and unadjusted modified Poisson regression.

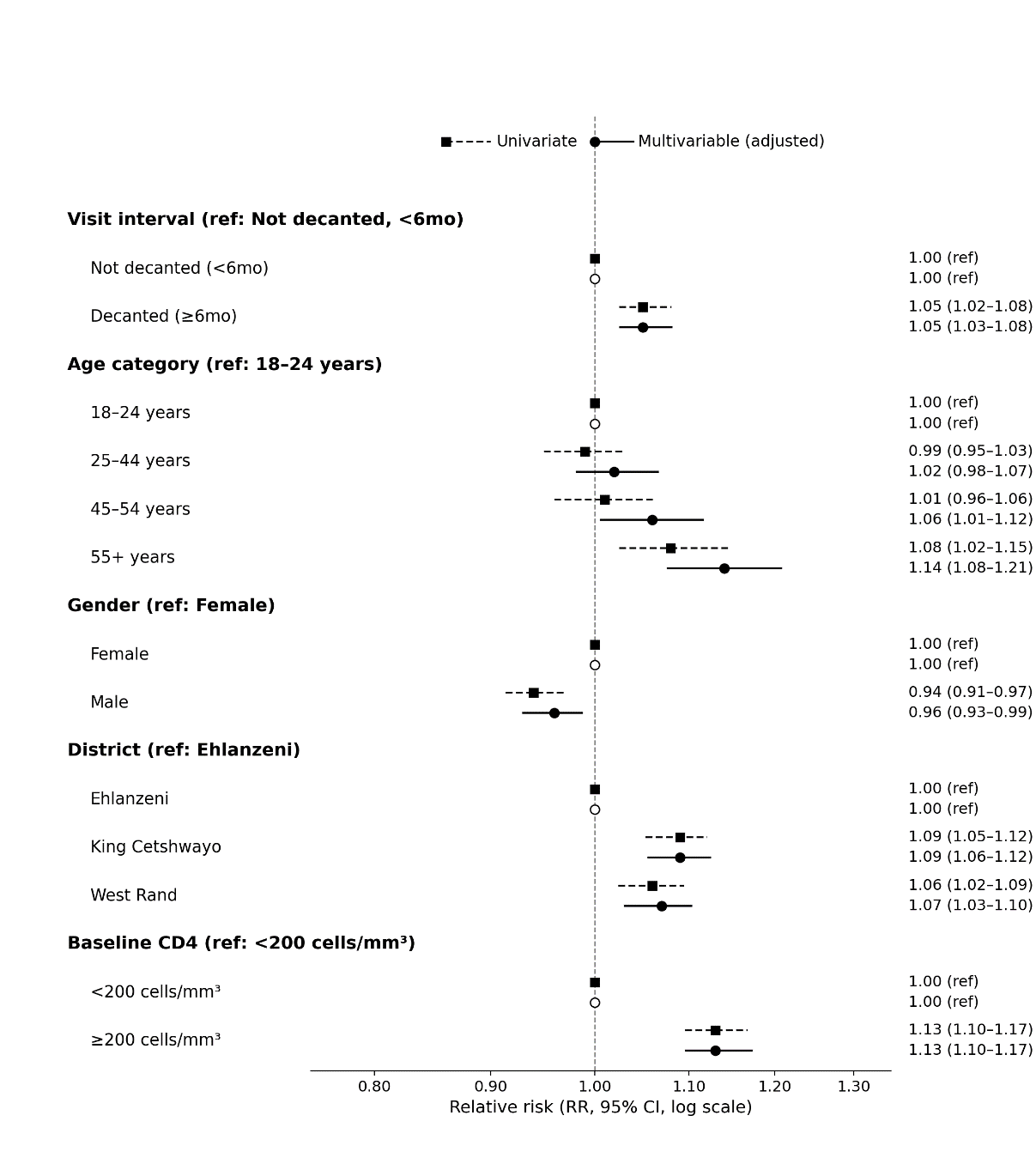

### Supplementary Figure 4. Number of clinic visits in first year on ART, year of initiation, 2022-2024

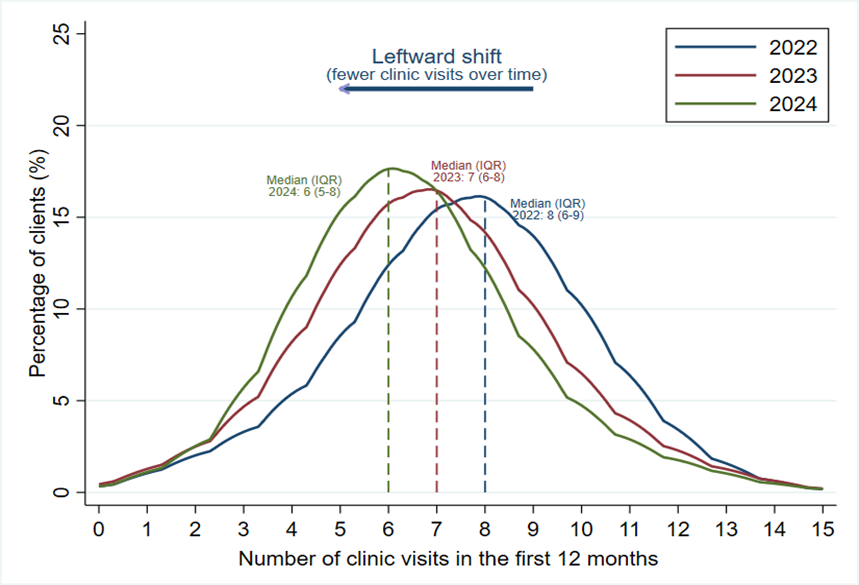
